# Evaluating discharges and readmissions using a COVID Virtual Ward model: a retrospective data study assessing patient outcomes and the likely staffing commitment

**DOI:** 10.1101/2021.07.16.21260651

**Authors:** Suzy Gallier, Catherine Atkin, Vinay Reddy-Kolanu, Dhruv Parekh, Xiaoxu Zou, Felicity Evison, Simon Ball, Elizabeth Sapey

**Affiliations:** PIONEER Technical Director, Lead for Research Analytics Department of Health Informatics, University Hospitals Birmingham NHS Foundation Trust, Edgbaston, Birmingham, B15 2GW, UK.; Acute Medicine, Birmingham Acute Care Research Group, Institute of Inflammation and Ageing, University of Birmingham, Edgbaston, Birmingham, B15 2GW, UK.; Acute Medicine, University Hospitals Birmingham NHS Foundation Trust, Edgbaston, Birmingham, B15 2GW, UK.; A. Intensive Care Medicine, University Hospitals Birmingham NHS Foundation Trust, Edgbaston, Birmingham, B15 2GW, UK., B. Birmingham Acute Care Research Group, Institute of Inflammation and Ageing, University of Birmingham, B15 2TT, UK.; Research Analytics Department of Health Informatics, University Hospitals Birmingham NHS Foundation Trust, Edgbaston, Birmingham, B15 2GW, UK.; A. Chief Medical Officer, University Hospitals Birmingham NHS Foundation Trust, Edgbaston, Birmingham, B15 2GW, UK.,, B. Better Care Programme Director, Health Data Research UK; A. Director of PIONEER: Health Data Research UK (HDRUK) Health Data Research Hub for Acute Care, B. Birmingham Acute Care Research Group, Institute of Inflammation and Ageing, University of Birmingham, B15 2TT, UK, C. Acute Medicine, University Hospitals Birmingham NHS Foundation Trust, Edgbaston, Birmingham, B15 2GW, UK

**Keywords:** Virtual ward, discharge, readmissions, mortality, intensive care

## Abstract

**Background:** COVID-19 has placed a catastrophic burden on acute hospitals. In an attempt to reduce admissions and enable safe early discharge, a COVID virtual ward (CVW) care pathway has been supported by NHS England. This includes discharging people who meet objective criteria based on acuity scores and oxygen saturations, with pulse oximeters and daily phone calls for up to 14 days. Observational studies have reported the safety of this system, but without describing the outcomes from usual care.

**Methods:** A retrospective study using routinely collected health data from all adults with a confirmed positive severe acute respiratory syndrome coronavirus-2 (SARS-CoV-2) swab result between 1^st^ June 2020 and 31^st^ Jan 2021 who attended the Emergency Department or Acute Medical Unit at QEHB, which does not have a CVW service. Criteria for CVW were applied using data from the first 24 hours of presentation to hospital and subsequent health outcomes were included for 28 days, including re-presentation, re-admission, ITU escalation and death. Results were compared to reported studies based in secondary care.

**Results:** During the study period, 26,127 patients presented to QEHB hospital. 2301 had a positive SARS-CoV-2 swab. Of these, 1730 (75.2%) did not meet the criteria for the CVW and 571 (24.8%) did. Of the 571, 325 (56.9%) were discharged home within 24 hours and 246 (43.1%) were admitted for 24 hours or longer. Those admitted were older, with increased co-morbidities, 80.9% required hospital-supported acute therapies after the first 24 hours and 10.6% died. Of the 325 discharged, 44 were readmitted (13.5%), 30 (9.2%) with COVID-related symptoms, 5 (1.5%) required ITU and 1 patient (0.3%) died. These results were comparable to published studies with a CVW service.

**Discussion:** In the current study, discharging patients without a CVW did not confer a greater risk of re-presentation, re-admission, ITU escalation or death. The majority of patients who remained in hospital despite meeting the CVW criteria did so for the provision of treatments or acute assessments. It remains uncertain whether a CVW delivers improvements in hard outcomes, and further research is needed.

## Introduction

The global pandemic caused by SARS-CoV-2 continues to provide health challenges worldwide. During infection ‘waves’, affected areas experience a high number of hospital presentations and admissions(1), requiring significant reconfiguration of staff and health services to meet the demands for care.

On 13^th^ January 2021, National Health Service England (NHSE) published a document which provided a standard operating procedure for a ‘COVID Virtual Ward’ (CVW), with an aim of enabling the safe discharge of patients with COVID-19 from the hospital setting to the community early upon presentation to hospital, with daily contact and safety netting. NHSE recommended that all areas should pursue the roll out of a COVID virtual ward model to reduce pressures on acute hospital services, and that the CVW should be delivered by the acute hospital(2).

The recommended CVW model was as follows(2): Subject to completion of a satisfactory exercise test, patients with oxygen saturations of 95-100% and low NEWS2 (< 3) and improving clinical trajectories could be discharged to a COVID virtual ward where clinically appropriate. Patients with saturations of 93-94% with improving clinical trajectories (symptoms, signs, blood results, CXRs), could also be considered for the COVID virtual ward where clinically appropriate. Patients with oxygen saturations of 92% or lower or experiencing moderate/severe shortness of breath would generally be unsuitable for early supported discharge, unless the patient was stable and this represented their usual baseline oxygen saturation.

In the CVW model, patients are discharged home with an oximeter and asked to take three readings each day and to call a staffed hospital number immediately if they note a reading of less than 92%, or they should attend their nearest emergency department within an hour or call 999. The patient would be proactively contacted by phone every day (seven days a week, as they would be for a hospital-based ward round). At 14 days (or before if deemed clinically appropriate) patients would either be discharged from the CVW or receive a further clinical assessment if symptomatic. A friend or family member, or an NHS Volunteer Responder, would then return the oximeter for decontamination and reuse.

To deliver this model, the CVW telephone line requires staffing for at least 12 hours a day (8am–8pm) seven days a week with locally arranged provision of out of hours cover. These staff should be supervised by an experienced, clinically registered professional who is also responsible for making the proactive daily calls, i.e. virtual ward round. The CVW has a named medical consultant or senior trainee (ST3+ doctor), usually an acute or respiratory physician.

This or similar models have been applied in a number of care settings. In June 2020, an article reported that of 200 patients managed on a CVW in a UK secondary care hospital, 13% on all cases re-presented to hospital, and 10% of all cases were readmitted(3). In November 2020, a retrospective study of 273 COVID-19 was published assessing a similar system for patients but with 5 days of virtual follow up rather than the 14 days described by NHSE, and also the option for discharge without enrolment to the CVW for those at lowest risk(4). The authors describe an 11% readmission rate and 1 death. A multi-site mixed methods study assessing clinical sites which had implemented a similar service suggested the cost of implementing such a service was approximately £400 to £553 per patient(5).

These studies support the safety of the CVW system. However, there are implied assumptions; first that all patients would otherwise be admitted to hospital and second, that the provision of the CVW impacts positively on re-presentations (either reducing unnecessary readmissions or identifying where re-assessment is needed). Also, there are no descriptions of the patients who did not meet the CVW criteria, to understand the volume of patients who still required in patient care. Implementation of such a system as the CVW has an opportunity cost, as staff have to be redeployed from other clinical areas and the reliance on acute or respiratory physicians to deliver the CVW may re-direct care from high intensity clinical environments during pandemic waves. Traditionally these care pathways would be assessed in randomised control trials with health economic reviews. In the absence of such trials, understanding both the “natural history” of those discharged who meet the CVW criteria and the care burden which remains in the hospital is important to place the staffing needs of the CVW in context.

Birmingham is one of the most ethnically diverse cities in the UK with a high burden of COVID-19 cases and COVID-19 associated mortality in all the UK COVID-19 waves(6, 7). This study was conducted to understand the potential impact of a CVW, by assessing the outcomes and clinical pathways of those presenting to hospital who both met and did not meet the criteria for potentially being included in a CVW, and assessing the time needed to deliver such a system in a hospital trust with a high burden of COVID-19 presentations.

## Methods

This data study was supported by PIONEER, a Health Data Research Hub in Acute Care. All study activities complied with the ethical approvals provided by the East Midlands – Derby REC (reference: 20/EM/0158).

University Hospitals Birmingham NHS Foundation Trust (UHB), UK is one of the largest Trusts nationally, covering 4 NHS hospital sites, treating over 2.2 million patients per year and housing the largest single critical care unit (CCU) in Europe. The Queen Elizabeth Hospital Birmingham (QEHB) is the largest hospital within UHB. UHB saw the highest number of COVID admissions in the UK (>10,000 confirmed cases by March 2021) and the highest number of patients ventilated, with an expanded Intensive Care Unit (ICU) capacity of >200 beds.

### Study population

All adults with a confirmed positive severe acute respiratory syndrome coronavirus-2 (SARS-CoV-2) swab result between 1^st^ June 2020 and 31^st^ Jan 2021 who attended the Emergency Department or Acute Medical Unit at QEHB at the time of or up to two weeks following a positive SARS-CoV-2 swab test were included. COVID cases were confirmed following a nasopharyngeal and oropharyngeal swab in all cases(8) which were processed in accordance with NHS guidance within UHB NHS laboratories (9).

UHB has built and runs its own electronic health record (EHR) and was able to develop a structured electronic clerking proforma where all patients suspected of having COVID-19 could be identified on admission. For all patients, the results of the first positive swab were included but patient records were checked for subsequent positive swab results if associated with a subsequent admission. Mortality and (in those alive) patient admission status (discharged and alive, continued admission and alive) were assessed 28 days after the first positive swab result (the latest date to assess outcome being 28th Feb 2021).

### Data Collection and variable definitions

Patient demographics and clinical data were collected from the QEHB EHR. Clinician confirmed co-morbidities were available from the EHR, the summary primary care record (Your Care Connected) and from diagnostic codes derived from previous hospital episodes. The EHR encodes diagnoses using NHS Digital SNOMED CT browser(10) alongside and mapped on to ICD-10 codes(11) allowing inclusion of historically entered ICD10 codes.

English Indices of Deprivation scores were calculated using postcodes from the current data provided by the UK’s Ministry of Housing, Communities and Local Government (2019) Report(12). Ethnicity was self-reported by the patient or their family members on admission to hospital. Ethnicity was grouped as per national guidelines(13).

Results of exercise tests, physiological assessments, chest radiograph reports and acuity scores were used to determine the suitability for a COVID virtual ward. Trajectories were considered using the same parameters over the first 24 hours of admission. For the purposes of this study, patients with either oxygen saturations of 95-100% and low NEWS2 (< 3) and patients with saturations of 93-94% with improving clinical trajectories over the first 24 hours of admission (symptoms, signs, blood results, CXRs) were considered as being suitable for the COVID virtual ward. Trajectories had to include oxygen saturations improving to >94% on room air and no deterioration in NEWS2 scores to be considered improved enough to meet the CVW criteria. Oxygen saturations could be 92% or lower if these were compatible with baseline levels for the patients as demonstrated through previous monitoring.

All patients with a positive COVID-19 admitted during the time period were included with no exceptions. Decisions as to whether a patient met not did not meet the criteria for the CVW were made against objective parameters by one person clinically qualified person and ratified by another (both acute medicine trained), with both being blind to patient outcomes (including re-presentation, escalation to ITU care and death).

In all admitted cases, clinical note reviews were undertaken to assess whether any assessments or treatments were delivered which would preclude discharge during the COVID pandemic, including the need for intravenous therapies or social care support. Reasons included provision of treatments which could not be given at home – these included intravenous therapy, oxygen or other respiratory support, requiring new, treatment dose anti-coagulation (subcutaneously) or requiring increased social care.

### Outcomes

The primary outcome was re-presentation to the Emergency Department or Acute Medical Unit for any cause, or death within 28 days of discharge, as per national reporting(14). For those patients discharged from hospital, primary care records were checked and any patients admitted to hospital with COVID-19 and discharged who had died in the community within the censor period were noted. Those with an on-going admission were censored 28 days after a positive swab result.

### Statistics

Statistical analysis was performed using STATA (SE) version 15 (StataCorp LLC, Texas, USA). Baseline characteristics for the total population are presented as mean (standard deviation) or median (interquartile range) for continuous variables and as frequency (percentage) for categorical variables. Continuous variables were compared between data sets using Mann-Whitney U tests. Categorical variables were compared using Fisher exact and Chi-Square tests. Results were considered significant if the p-value was <0.05. There was no adjustment for multiple comparisons but exact p values are given.

## Result

In total, 2301 swab confirmed cases of COVD-19 were assessed in the 245day time period. Of note, 23,803 patients who were COVID negative were assessed in the same period, of which 2827 (11.9%) were discharged and 20740 (87.1%) admitted. Figure 1 shows a diagram of overall patient numbers across categories.

**Figure 1.**
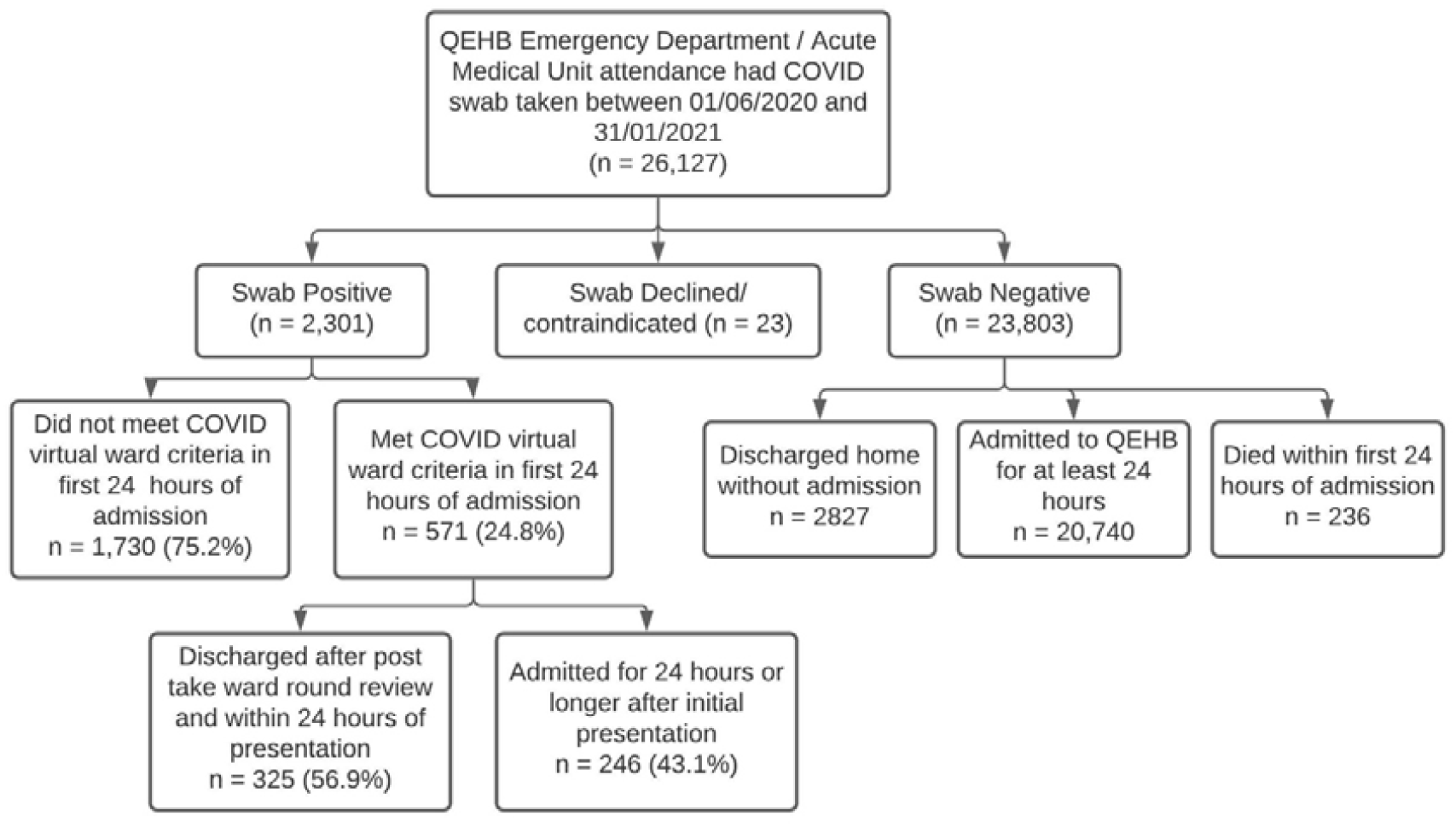
A modified consort diagram of patients presenting to the Emergency Department of Acute Medical Unit at QEHB. Presentations were all presentations for the time period stated. Only those with a confirmed positive PCR SARS-CoV-2 Swab were included in pathway mapping. Those with no swab results (23 individuals where swabs were declined or contraindicated due to facial injury) were not included in the pathway analysis. Patients were divided into those meeting the COVID Virtual Ward criteria as described in the methods.

### Patient demographics

The overall demographics of all patients presenting with COVID-19 during the time period who did or did not meet the CVW criteria are given in table 1, irrespective of whether they were discharged or not. Table 2 compares the demographics of patients who met the CVW criteria, comparing those who were discharged within 24 hours of presentation and those who were admitted for ≥24 hours. In general, patients who remained in-patients were more likely to be older, with 14.5% of patients aged >65 years in the discharged group, compared to 44.8% in the admitted group. Those admitted were more likely to have significant co-morbidities, including dementia, stroke, active malignancies and heart disease.

**Table 1.** A comparison of demographics between swab positive patients who met or did not meet the COVID virtual ward criteria between 1^st^ June 2020 and 31st January 2021, irrespective of admission or discharge. Data is number (percentage) unless otherwise stated. Ethnicity was self-reported (see Methods). English Indices of deprivation (IMD) were calculated using postcode. Diabetes includes type 1 and type 2 diabetes. COPD= Chronic Obstructive Pulmonary Disease. Patients could (and often did) have more than one co-morbid condition^*^

|  |  | Met the virtual<br>COVID ward criteria | Did not meet<br>virtual COVID ward<br>criteria | p-value |
| --- | --- | --- | --- | --- |
|  | Overall Count | 571 (24.8%) | 1730 (75.2%) |  |
| Sex | Female | 309 (54.1%) | 803 (46.4%) | **p = 0.001** |
|  | Male | 262 (45.9%) | 926 (53.5%) |  |
| Age | Median (IQR) | 51 (37-68) | 62 (50-78) | **p<0.001** |
|  | 50 and Under | 285 (49.9%) | 445 (25.7%) |  |
|  | 51-65 | 129 (22.6%) | 525 (30.3%) |  |
|  | 66-75 | 52 (9.1%) | 267 (15.4%) |  |
|  | 76-85 | 54 (9.5%) | 280 (16.2%) |  |
|  | Over 85 | 51 (8.9%) | 213 (12.3%) |  |
| Ethnicity | White | 238 (41.7%) | 823 (47.6%) | p = 0.124 |
|  | Black | 30 (5.3%) | 79 (4.6%) |  |
|  | South Asian | 141 (24.7%) | 425 (24.6%) |  |
|  | Mixed | 14 (2.5%) | 34 (2%) |  |
|  | Chinese | 3 (0.5%) | 9 (0.5%) |  |
|  | Any Other Ethnic Group | 18 (3.2%) | 61 (3.5%) |  |
|  | Unknown or declined to self-report | 127 (22.2%) | 299 (17.3%) |  |
| IMD Quintile | 1 (Most Deprived) | 308 (53.9%) | 943 (54.5%) | p = 0.765 |
|  | 2 | 127 (22.2%) | 351 (20.3%) |  |
|  | 3 | 79 (13.8%) | 266 (15.4%) |  |
|  | 4 | 40 (7%) | 108 (6.2%) |  |
|  | 5 (Least Deprived) | 16 (2.8%) | 60 (3.5%) |  |
|  | Unknown | 1 (0.2%) | 2 (0.1%) |  |
| Co-morbidities* | Chronic Kidney Disease | 69 (12.1%) | 259 (15%) | p = 0.087 |
|  | Dementia | 38 (6.7%) | 170 (9.8%) | **p = 0.022** |
|  | Interstitial Lung Disease | 5 (0.9%) | 29 (1.7%) | p = 0.169 |
|  | Stroke or cerebrovascular | 65 (11.4%) | 243 (14%) | p = 0.105 |
|  | Ischaemic heart disease | 108 (18.9%) | 364 (21%) | p = 0.275 |
|  | Asthma | 101 (17.7%) | 343 (19.8%) | p = 0.262 |
|  | Hypertension | 205 (35.9%) | 903 (52.2%) | **p < 0.001** |
|  | Diabetes | 132 (23.1%) | 586 (33.9%) | **p < 0.001** |
|  | Any active malignancy | 91 (15.9%) | 334 (19.3%) | p = 0.072 |
|  | COPD | 28 (4.9%) | 224 (12.9%) | **p < 0.001** |
|  | Atrial fibrillation | 63 (11%) | 236 (13.6%) | p = 0.108 |
| BMI | Underweight | 13 (2.3%) | 40 (2.3%) | **p = 0.003** |
|  | Normal weight | 93 (16.3%) | 338 (19.5%) |  |
|  | Overweight | 91 (15.9%) | 463 (26.8%) |  |
|  | Obese | 78 (13.7%) | 508 (29.4%) |  |
|  | Morbid obesity | 21 (3.7%) | 136 (7.9%) |  |

**Table 2.** A comparison of demographics between swab positive patients who met the COVID virtual ward criteria between 1^st^ June 2020 and 31^st^ Jan 2021 who were either discharged within 24 hours of attendance or admitted to hospital for > 24 hours. Data is number (percentage) unless otherwise stated. Ethnicity was self-reported (see Methods). English Indices of deprivation (IMD) were calculated using postcode. Diabetes includes type 1 and type 2 diabetes. COPD= Chronic Obstructive Pulmonary Disease. Patients could (and often did) have more than one co-morbid condition

|  |  | Discharged within 24 hours | Admitted for > 24 hours | p-value |
| --- | --- | --- | --- | --- |
|  | Overall Count | 325 (56.9%) | 246 (43.1%) |  |
| Sex | Female | 185 (56.9%) | 124 (50.4%) | p = 0.122 |
|  | Male | 140 (43.1%) | 122 (49.6%) |  |
| Age | Median (IQR) | 45 (34-57) | 63 (44-80) | **p < 0.001** |
|  | 50 and Under | 202 (62.2%) | 83 (33.7%) |  |
|  | 51-65 | 76 (23.4%) | 53 (21.5%) |  |
|  | 66-75 | 25 (7.7%) | 27 (11%) |  |
|  | 76-85 | 12 (3.7%) | 42 (17.1%) |  |
|  | Over 85 | 10 (3.1%) | 41 (16.7%) |  |
| Ethnicity | White | 114 (35.1%) | 124 (50.4%) | **p = 0.010** |
|  | Black | 16 (4.9%) | 14 (5.7%) |  |
|  | South Asian | 90 (27.7%) | 51 (20.7%) |  |
|  | Mixed | 10 (3.1%) | 4 (1.6%) |  |
|  | Chinese | 1 (0.3%) | 2 (0.8%) |  |
|  | Any Other Ethnic Group | 13 (4%) | 5 (2%) |  |
|  | Unknown | 81 (24.9%) | 46 (18.7%) |  |
| IMD Quintile | 1 (Most Deprived) | 175 (53.8%) | 133 (54.1%) | **p = 0.012** |
|  | 2 | 76 (23.4%) | 51 (20.7%) |  |
|  | 3 | 48 (14.8%) | 31 (12.6%) |  |
|  | 4 | 23 (7.1%) | 17 (6.9%) |  |
|  | 5 (Least Deprived) | 2 (0.6%) | 14 (5.7%) |  |
|  | Unknown | 1 (0.3%) | 0 (0%) |  |
| Co-morbidities* | Chronic Kidney Disease | 14 (4.3%) | 55 (22.4%) | **p < 0.001** |
|  | Dementia | 6 (1.8%) | 32 (13%) | **p < 0.001** |
|  | Interstitial Lung Disease | 2 (0.6%) | 3 (1.2%) | p = 0.443 |
|  | Stroke or cerebrovascular | 9 (2.8%) | 56 (22.8%) | **p < 0.001** |
|  | Ischaemic heart disease | 32 (9.8%) | 76 (30.9%) | **p < 0.001** |
|  | Asthma | 50 (15.4%) | 51 (20.7%) | **p = 0.097** |
|  | Hypertension | 66 (20.3%) | 139 (56.5%) | **p < 0.001** |
|  | Diabetes | 41 (12.6%) | 91 (37%) | **p < 0.001** |
|  | Any active malignancy | 20 (6.2%) | 71 (28.9%) | **p < 0.001** |
|  | COPD | 13 (4%) | 15 (6.1%) | p = 0.250 |
|  | Atrial fibrillation | 15 (4.6%) | 48 (19.5%) | **p < 0.001** |
| BMI | Underweight | 2 (0.6%) | 11 (4.5%) | p = 0.436 |
|  | Normal weight | 19 (5.8%) | 74 (30.1%) |  |
|  | Overweight | 22 (6.8%) | 69 (28%) |  |
|  | Obese | 21 (6.5%) | 57 (23.2%) |  |
|  | Morbid obesity | 8 (2.5%) | 13 (5.3%) |  |

### Outcomes for those discharged from hospital after initial review

Outcomes for the 325 swab positive patients who met the COVID virtual ward criteria between 1st June 2020 and 31^st^ Jan 2021 and were discharged within 24 hours were assessed for 28 days after initial presentation. 281 patients (86.4%) were not readmitted and were still alive at 28 days post initial presentation. 44 patients (13.5%) re-presented within 28 days to QEH, 30 of these (68.2% in total or 9.2% of the total cohort of 325 patients) were for COVID-related symptoms. The remaining 14 presented with a diverse list of conditions, listed in Table 3. Overall, the median length of time at home prior to re-presentation was 2.5 days (IQR 1.5 – 5 days).

**Table 3.** The list of reasons COVID-positive patients re-presented to hospital within 28 days after discharge. Data is number (percentage). Data was collected via routine clinical coding but individual notes were checked to confirm medical reason for re-presentation by a consultant physician.

| Readmission diagnosis | No of Pts | % of Total |
| --- | --- | --- |
| COVID- associated re-presentation | 30 | 68.20% |
| Poisoning | 3 | 6.82% |
| Musculoskeletal / limb pain | 3 | 6.82% |
| Fractured bone | 2 | 4.55% |
| Congestive heart failure | 1 | 2.30% |
| Epilepsy, unspecified | 1 | 2.30% |
| Headache | 1 | 2.30% |
| Pilonidal cyst with abscess | 1 | 2.30% |
| Pulmonary embolism | 1 | 2.30% |
| Syncope and collapse | 1 | 2.30% |
| Total | 44 |  |

Nine (20.5%) of the 44 patients were discharged within 24 hours of this second presentation. Of the COVID-associated re-presentations, the overall median length of stay in hospital was 4 days (IQR 1 – 7.5 days). Of the 44 re-presenting patients, 5 (11.4%) were transferred to ITU. 1 (2.3%) patient died and 43 (97.7%) patients survived to at least 28 days post initial COVID-19 presentation.

### Outcomes for those who were admitted to hospital after initial review

Treatment pathways and outcomes for the 246 swab positive patients who met the COVID virtual ward criteria between 1st June 2020 and 31^st^ Jan 2021 and were admitted to hospital for more than 24 hours were assessed for 28 days after the initial presentation. 191 patients (77.6%) received an intravenous infusion including fluids or antibiotics after the first 24 hours of admission. 14 patients (5.7%) received therapeutic dose anti-coagulation. 94 patients (38.2%) required oxygen therapy after the initial 24-hour period. 199 patients (80.9%) received either intravenous treatment, supplemental oxygen or required a community care hospital assessment. 47 patients did not require intravenous treatments, supplemental oxygen or a community hospital assessment.

For these 246 patients, the median length of stay in hospital was 4 days (IQR 2 – 9 days). 9 patients (3.7%) were transferred to intensive care. 26 patients (10.6%) died within 28 days of presenting to the emergency department and 220 (89.4%) remained alive during this time. Of those 26 who died, 18 died during the in-patient stay.

Of those who survived to discharge, 215 (87.4%) patients were discharged to their own home or to their usual care home (204 patients were discharged to a private dwelling and 11 pts to their usual care home). 12 (4.8%) were discharged to a community healthcare hospital. For the 47 patients who did not require intravenous therapies or oxygen or community care review, the median length of stay was 34 hours (IQR 28 – 40 hours) and none of these patients were readmitted during the follow up period. They were younger (median age 47 years (35-59), p=0.002) but with co-morbidities. A notes review revealed that these patients were not reviewed by an Acute Medical or Respiratory consultant within the first 24 hours of their admission.

### Comparing reported outcomes

Comparing the outcomes of patients who had been discharged without the CVW in the current study, to the description of the study from Thornton(3), there were no differences in the percentage of patients re-presenting. Also, there were also no differences in the percentage of patients re-presenting between Nunan et al (4) with a slightly different version of the CVW and UHB with no CVW. See table 4.

**Table 4:** Comparing re-presentations to hospital with and without the CVW. Numbers are those who either were discharged to the CVW (Thornton and Nunan) or met the criteria for the CVW and were discharged from UHB without a CVW in place. Re-presentation is defined as reattendance at the Emergency Department or acute medical unit. Figures are compared using Fishers Exact tests ^**^ = comparing reported Nunan at el(4) and UHB re-presentation rates.

|  | Total numbers | Re-presented to hospital | Did not re-present to hospital | p-value |
| --- | --- | --- | --- | --- |
| Thornton(3) | 200 | 26 | 174 | $p = 0.895^*$ |
| UHB | 325 | 44 | 281 |  |
| Nunan et al(4) | 273 | 31 | 242 | $p = 0.458^{**}$ |

Where reported, there were no differences in rates of admission to ITU in patients who re-presented to hospital while on the CVW in Nunan et al (4) compared to the UHB data; 2 were admitted to ITU from 31 re-presentations (4) versus 5 admissions to ITU at UHB from 44 re-presentations, Fisher’s exact test, p=0.693). There was one death in patients who re-presented on the CVW in Nunan et al (4) and one death in patients who re-presented in the current study, with again no statistical differences in event numbers (Fisher Exact, p = 0.999).

### Time required for service provision

If modelling virtual care requirements of the 325 who were sent home within 24 hours, and assuming 14 days of follow up (as per NHSE guidance) and a steady admission rate during the assessed time period, 4550 telephone calls would be required in the 245 days of the study period + 14 days to allow for follow up (259 days), an average of 18 calls per day. If each telephone contact required 25 minutes (5 minutes preparation/ 5 minutes note making and 15 minutes conversation with the patient), the daily workload would be 450 minutes/7.5 hours each day (without break), or 52.5 hours per week, for every week of the study period. While it is unlikely that all patients who met the CVW criteria within the first 24 hours of presentation would have been deemed medically fit for discharge to the virtual ward, the same modelling for the 571 cohort who met the CVW criteria would require 7994 telephone calls over the study period plus follow up period of 14 days, which equates to 31 calls per day, taking 775 minutes or 13 hours per day. This excludes the provision of a manned telephone service that patients could call if concerned. This would require at least 2 or 3 staff members each day, as well as senior medical oversight and out of hours cover.

## Discussion

The NHSE CVW was developed to facilitate the care of COVID-19 patients who had presented to hospital, but who could be cared for at home. Thrice daily oxygenation saturations and daily contact by trained hospital staff were provided for appropriate safety netting. The hope for the service is that it would free hospital beds, enable staff to focus on the most unwell patients with COVID-19, while providing a safe clinical service for patients.

To date, the CVW has not been evaluated in a gold standard, robust randomised clinical trial. Instead, CVWs have been reported in a series of observational studies and summarised in a recent systematic review(15). These studies present some evidence of the safety of the CVW service, but have not described the burden of COVID where criteria were not met, nor described the natural history of discharged patients without this service, to enable a comparison of opportunity costs as well as gains.

This study presents retrospectively analysed, routinely collected health data for all patients admitted to QEHB with a positive COVID-19 swab result. Patients were assessed as to whether they met the criteria for the CVW parameters as presented by NHSE(2) either on initial presentation or during the first 24 hours, using time-stamped data available in the clinical record.

Of note, 75.2% (1730) of patients presenting to hospital during this period did not meet the CVW criteria, suggesting that during a wave of COVID-19, the CVW would only be suitable for a proportion of patients. In the 24.8% (571) who did meet the CVW criteria, 325 (56.9%) patients were discharged, and of those discharged, only 13% re-presented and 10% were admitted, percentages which are the same as reported in those discharged on the CVW, suggesting the CVW may not reduce re-presentation or re-admission rates. Further, the proportion of re-presenting patients who required ITU care or who died were the same as those reported in one CVW observational study(4), suggesting the CVW may not be safer in practice than discharge without the CVW. This study does not include a health economic analysis of service provision, but during COVID-19 waves, the CVW requires substantial staffing to operate. It is unclear whether there is the capacity to redeploy these skilled staff members from caring for the 75% of patients who did not meet CVW criteria to the 25%, especially if this does not confer improvements in hard outcomes such as mortality, ITU provision or re-presentation and admission. Previous studies have reported patient satisfaction with the CVW, reducing the potential anxiety of being discharged(4). The current study does not assess this important factor.

Of note, a number of studies have reported remote monitoring for COVID-19 patients either initially in the community for positive patients as part of a triage system with lower presentations to hospital (5), such as 4% (16), which probably reflects the screening of milder cases, less likely to present to hospital in the current study. However, where focusing on a secondary care delivered CVW, a re-presentation rate of approximately 13% has been reported(17).

This study has limitations. First, by comparing outcomes across studies, there is an assumption that patient populations including demography, burden of disease and impact of COVID-19, are similar across published studies. Most studies to date have not presented in depth demography and therefore these direct comparisons should be reviewed with caution. Second, in the current paper, the objective criteria for the CVW were applied using the granular electronic health record at QEHB without the benefit of seeing the patient or considering the time of the initial presentation, and all these factors are important when making the decision to admit or discharge. Third, it is possible that more of the patients who met the CVW criteria might have been discharged were the CVW in place. Fourth, when assessing staffing needs it has been assumed that all patients will require follow up for 14 days, as described in the NHSE SOP. It is highly likely that some would require much less follow up prior to discharge. Fifth, this is not a formal health economic review, which would form part of the evaluation of any new service.

The study has significant strengths. Decisions as to whether the patient met the CVW criteria were made independently by two researchers, without knowledge of outcomes, thus reducing bias. All patients were included in the study, reducing the population bias which can hinder consented studies. As data includes all medications and electronic noting, reasons for continued admission could be assessed.

In summary, this study reports similar percentage re-presentations, admissions, escalations to ITU and deaths in patients who were discharged without a CVW compared to studies where a CVW was implemented. The study also highlights that only 25% of COVID-19 admissions were suitable for the CVW based on objective criteria, and the potential intensity of work created by implementing a CVW. The study begins to explore the potential opportunity costs of the CVW system, but does not consider patient factors, such as the reassurance potentially given by discharge to a CVW setting.

Any new care pathways or initiatives have opportunity gains and costs, and in usual times, processes are often assessed using rigorous randomised control trials which often include quantitative, qualitative and health economic analyses. While the CVW was established to enhance patient safety and reduce unneeded hospitalisation, it is important to assess whether the service achieves this compares to usual care. More research is needed before this service is fully implemented.

## Data Availability

Data Access: Data from this study is available from PIONEER, the Health Data Hub in Acute care, in accordance with Hub processes. Contact for more details.

## Acknowledgements

This work was supported by PIONEER, the Health Data Research Hub in acute care, and the Better Care Programme, both funded by Health Data Research UK. This work uses data provided by patients and collected by the NHS as part of their care and support. We would like to acknowledge the contribution of all staff, key workers, patients and the community who have supported our hospitals and the wider NHS at this time.

## Author contributions

C.A, V.R-K, D.P assisted with clinical data insights, F.E, X.Z and S.G analysed the data. S.G, S.B and E.S designed the study and wrote the manuscript. All authors approved the final manuscript.

## Data Access

Data from this study is available from PIONEER, the Health Data Hub in Acute care, in accordance with Hub processes. Contact for more details.

